# Antimicrobial use in COVID-19 patients in the first phase of the SARS-CoV-2 pandemic: Rapid review and evidence synthesis

**DOI:** 10.1101/2021.02.18.21251932

**Authors:** Wenjuan Cong, Ak Narayan Poudel, Nour Alhusein, Hexing Wang, Guiqing Yao, Helen Lambert

## Abstract

**Background:** As the numbers of people with COVID-19 continue to increase globally, concerns have been raised regarding the widespread use of antibiotics for the treatment of COVID-19 patients and its consequences for antimicrobial resistance during the pandemic and beyond. The scale and determinants of antibiotic use in the early phase of the pandemic, and whether antibiotic prescribing is beneficial to treatment effectiveness in COVID-19 patients, are still unknown. Unwarranted treatment of this viral infection with antibiotics may exacerbate the problem of antibiotic resistance, while antibiotic resistance may render presumptive treatment of secondary infections in COVID-19 patients ineffective.

**Methods:** This rapid review was undertaken to identify studies reporting antimicrobial use in the treatment of hospitalised COVID-19 patients. The review was conducted to comply with PRISMA guidelines for Scoping Reviews (http://www.prisma-statement.org/Extensions/ScopingReviews) and the protocol was registered with the Open Science Framework (OSF): http://osf.io/vp6t5. The following databases: Web of Science, EMBASE, PubMed, CNKI & VIP were searched to identify the relevant studies from 1 Dec 2019 up to 15 June 2020; no limits were set on the language or the country where studies were conducted. The search terms used were: ((“Covid-19” or “SARS-CoV-2” or “Coronavirus disease 2019” or “severe acute respiratory syndrome coronavirus-2”) and ((“antibiotic prescribing” or “antibiotic use” or “antibiotic*”) or “antimicrobial *” or “antimicrobial therapy” or “antimicrobial resistance” or “antimicrobial stewardship”)). A total of 1216 records were identified through database searching and 118 clinical studies met the inclusion criteria and were taken into data extraction. A bespoke data extraction form was developed and validated through two independent, duplicate extraction of data from five Records. As all the included studies were descriptive in nature, we conducted descriptive synthesis of data and reported pooled estimates such as mean, percentage and frequency. We created a series of scenarios to capture the range of rationales for antibiotic prescribing presented in the included studies.

**Results:** Our results show that during the early phase of the pandemic, 8501 out of 10 329 COVID-19 patients (82·3%) were prescribed antibiotics; antibiotics were prescribed for COVID-19 patients regardless of reported severity, with a similar mean antibiotic prescribing rate between patients with severe or critical illness (75·4%) and patients with mild or moderate illness (75·1%). The top five frequently prescribed antibiotics for hospitalised COVID-19 patients were azithromycin (28·0 % of studies), ceftriaxone (17·8%), moxifloxacin (14·4%), meropenem (14·4%) and piperacillin/tazobactam (12·7%). The proportion of patients prescribed antibiotics without clinical justification was 51·5% vs 41·9 % for patients with mild or moderate illness and those with severe or critical illness respectively. Comparison of patients who were provided antibiotics with a clinical justification with those who were given antibiotics without clinical justification showed lower mortality rates (9·5% vs 13·1%), higher discharge rates (80·9% vs 69·3%) and shorter length of hospital of stay (9·3 days vs 12·2 days). Only 9·7% of patients in our included studies were reported to have secondary infections.

**Conclusions:** Antibiotics were prescribed indiscriminately for hospitalised COVID-19 patients regardless of severity of illness during the early phase of the pandemic. COVID-19 related concerns and lack of knowledge drove a large proportion of antibiotic use without specific clinical justification. Although we are still in the midst of the pandemic, the goals of antimicrobial stewardship should remain unchanged for the treatment of COVID-19 patients.

## Introduction

Antimicrobial resistance (AMR) kills an estimated 700 000 people every year.^1^ Without intervention, the current trajectory predicts a gloomy figure of 10 million fatalities by 2050.^2^ The SARS-CoV-2 pandemic foreshadows the crisis of living with an infectious disease for which there is no treatment and the damaging consequences to our health systems and economies. At the beginning of the pandemic, with the panic of facing the unknown, many existing medicines were repurposed to treat the virus. This included widespread use of antibiotics in treatment.^3-7^ For example, in a multi-hospital cohort study in the USA, 56·6% of 1705 patients were prescribed early empiric antibacterial therapy, of which only 3·5% were confirmed to have bacterial infection.^5^ Two systematic reviews found that, of the patients reported in the included studies, 72·0% received antibiotics, 14·3% suffered a secondary bacterial infection.^4,7^ The low proportion of COVID-19 patients having coinfection or secondary infection in these studies is consistent with other findings. For example, in Italy, from the 16 654 patients who died of COVID-19, only 11% were reported to have a secondary bacterial infection (data as of April 09, 2020).^8^ In Spain, of 989 consecutive patients with COVID-19, 72 (7·2%) had 88 other microbiologically confirmed infections: 74 were bacterial, seven fungal and seven viral.^9^

Overall, the pandemic may be accelerating the threat of antimicrobial resistance (AMR) due to the increased use of antibiotics, increased exposure to hospital environments and invasive procedures used in COVID-19 treatment, while evidence for the benefits of antimicrobial use in such patients is limited. Many AMR experts have raised their concerns around the safety of using antibiotics in COVID-19 patients and called for strengthening antimicrobial stewardship (AMS) programs in the time of COVID-19. ^8, 10, 11, 12^-^14^ For example, the increased use of empirical antibiotics treatment increases the risks of *Clostridioides difficile* infection in COVID-19 patients and the emergence of multidrug resistant organism.^15,16^ Additionally, some have started to question the possible accumulation of inflammatory properties of some administrated medicines which might have contributed to inducing an inflammatory storm in COVID-19 patients.^17-19^

Guidelines have started to emerge around the use of antimicrobials in COVID-19 patients. For example, WHO guidelines recommend no antibiotic therapy or prophylaxis for patients with mild or moderate COVID-19 unless signs and symptoms of a bacterial infection exist. For severe COVID-19, a daily assessment for de-escalation of antimicrobial treatment is recommended. For elderly patients and children under five with moderate COVID-19, WHO recommends use of antibiotics categorised in the WHO access list of medicines such as co-amoxicillin.^10,20^ Other guidelines such as those of the Dutch Working Party on Antibiotic Policy^16^ and the Scottish Antimicrobial Prescribing Group^21^ both advise avoiding routine antibiotic use in suspected COVID-19, the importance of obtaining sputum and blood samples as well as urinary antigen testing upon admission, and a cautious antibiotic treatment of short duration of five days in patients of COVID-19 when there is a clinical suspicion of secondary bacterial infection.

There is an urgent need for further research and guidance in this field, from producing evidence-based guidelines, ^16^ reassessing biomarkers for antimicrobial stewardship in COVID-19 patients,^22^ understanding drivers, benefits and disbenefits of antibiotic use and assessing the wider impact of the pandemic on antimicrobial use (AMU) and AMR. In this scoping review, we aim to: add to the research evidence on prevalence and patterns of antimicrobial use in the treatment of COVID-19 patients; identify the most commonly used antibiotics in the treatment of hospitalised COVID-19 patients worldwide; identify the clinical scenarios associated with AMU; and to explore any impact on patient outcomes.

## Method

This rapid review was undertaken to identify, synthesise and analyse findings from studies that reported antibiotic use in the treatment of COVID-19 patients. The review was conducted to comply with PRISMA guidelines for Scoping Reviews (http://www.prisma-statement.org/Extensions/ScopingReviews);^23^ and the protocol has been registered with the Open Science Framework (OSF): http://osf.io/vp6t5. We selected a scoping rather than systematic review approach in order to maximise data inclusion from a wide range of study types.

### Search strategy

The following databases: Web of Science, EMBASE, PubMed and two Chinese academic databases (CNKI & VIP) were searched to identify relevant studies from 1 Dec 2019 up to 15 June 2020; no limits were set on the country where study was conducted, and we excluded any studies not available in English or Chinese. The search terms were: ((“COVID-19” or “SARS-CoV-2” or “Coronavirus disease 2019” or “severe acute respiratory syndrome coronavirus-2”) and ((“antibiotic prescribing” or “antibiotic use” or “antibiotic*”) or “antimicrobial *” or “antimicrobial therapy” or “antimicrobial resistance” or “antimicrobial stewardship”)).

### Web of Science

All Fields = (COVID-19 and antibiotic*) or (SARS-CoV-2 and antibiotic*) or (Coronavirus disease 2019 and antibiotic*) or (severe acute respiratory syndrome coronavirus-2 and antibiotic*)

All Fields = (COVID-19 and antimicrobial*) or (SARS-CoV-2 and antimicrobial*) or (Coronavirus disease 2019 and antimicrobial*) or (severe acute respiratory syndrome coronavirus-2 and antimicrobial*)

### PubMed

(COVID-19 or SARS-CoV-2 or Coronavirus disease 2019 or severe acute respiratory syndrome coronavirus-2) and antimicrobial* (COVID-19 or SARS-CoV-2 or Coronavirus disease 2019 or severe acute respiratory syndrome coronavirus-2) and antibiotic*

### Embase

(antibiotic* or antibiotic prescribing or antimicrobial resistance or antibacterial*) and (severe acute respiratory syndrome coronavirus-2 or COVID-19 or SARS-CoV-2 or Coronavirus disease 2019).

### CNKI & VIP

新型冠状病毒或新冠肺炎或 2019 冠状病毒或 COVID-19; 抗生素使用或抗菌药物使用或抗菌药物管理或抗生素耐药性

### Inclusion, exclusion criteria and study selection process

Articles fulfilling the following criteria were considered for inclusion in the review. Full-text articles only were included.

#### Inclusion criteria

1. All types of clinical studies (randomised control trial (RCT), cohort, case report including case series, other observational & descriptive studies) about the use of antibiotics to treat patients with COVID-19.
2. Studies reporting patients diagnosed with COVID-19 and receiving antibiotic treatment, without restrictions on age, race, gender, geographical location or setting.
3. Studies which had only mentioned antibiotic treatment without specifying the types of antibiotics or reporting treatment outcomes.

#### Exclusion Criteria

1. Animal studies, *in vitro* experiments, in silico screening/drug modelling, molecular mechanism and other aspects of COVID-19 research where not related to or mentioned antibiotic use (ABU)
2. Conference abstracts
3. Commentaries & editorial letters not reporting ABU
4. Literature review not reporting ABU
5. Trial protocol
6. Case report & case series not reporting ABU
7. Full-text articles not available in English or Chinese

Titles and abstracts were screened initially, and full texts were retrieved of articles which appeared to fulfil the inclusion criteria. Two independent, duplicate screenings were undertaken by WC & NA for all search results. Disagreement was resolved by consensus. Additional studies were identified by searching the reference lists of retrieved articles and the authors’ reference collections.

### Data extraction

A bespoke data extraction form was developed and validated through two independent, duplicate extraction of data from five relevant studies. Data extraction for antibiotic use in the treatment of COVID-19 patients included: publication details, region, type of COVID-19 patients, age, gender, number of patients reported, study type (case report including case series, RCT, cohort, other descriptive studies), type of patients (mild or moderate, severe or critical), antimicrobial prescribing rate (overall and for different types of patient, details of antibiotics prescribed if reported, antibiotic prescribing scenarios (under which circumstance antibiotic was prescribed), whether antibiotic treatment had complied with AMS practice (yes, no or not sure), mean length of hospital stay, discharge rate, and mortality rate.

Data extraction for COVID-19 patients with secondary bacteria or fungal infections included: publication details, region, gender, reported number of patients with co-infections, type of patients with co-infections (severe or critical ill), type of co-infections, details of antibiotic used for the treatment of co-infections, length of stay, discharge rate, mortality rate of patients with co-infections, gender, age and underlying health conditions of patient with co-infections if reported.

### Data synthesis and analysis

We looked at the overall antibiotic prescribing rate, scenarios of antibiotics prescribing, types of antibiotic use and health outcomes of patients. In this review, data extraction and analysis were performed in Microsoft Excel. We conducted descriptive synthesis, taking into account of the sample size of each study (i.e. weighted mean) while calculating pooled estimates of outcome variables (i.e. antibiotics prescribing %, mortality rate, discharge rate, length of stay). We also conducted subgroup analyses of COVID-19 patients who were given antibiotics during hospital admission, most frequently used antibiotics for these patients, mean percentage of clinically justifiable antibiotic use, most frequently used medicines (other than antibiotics) for COVID-19 patients, proportion of COVID-19 patients having underlying health conditions, mean length of stay in hospital, mean discharge rate and mean mortality rate.

As described above, we also investigated proportion of patients experiencing co-infections (fungal, bacterial or other), most frequently used antibiotics for the treatment of co-infections, mean length of stay for patients with co-infections, mean discharge rate and mean mortality rate for the patients with co-infections and proportion of patients with co-infection having underlying health conditions. We present these results by age and gender of patients.

## Results

### Study selection

A total of 1216 records were identified through database searching. After duplicates were removed and irrelevant records such as animal studies, *in vitro* experiments, in silico screening/drug modelling, molecular mechanism and other aspects of COVID-19 research not related to clinical treatment of COVID-19 patients were excluded, 223 records were screened for eligibility. A further 113 were excluded that did not report either antimicrobial use in the treatment or patients with co-infections (113/223, 51%). The remaining 110 articles and an additional 8 articles that were identified by searching the reference lists of the retrieved articles and the authors’ reference collections, led to a total of 118 full-text articles being included for review. (Fig 1)

**Fig 1.**
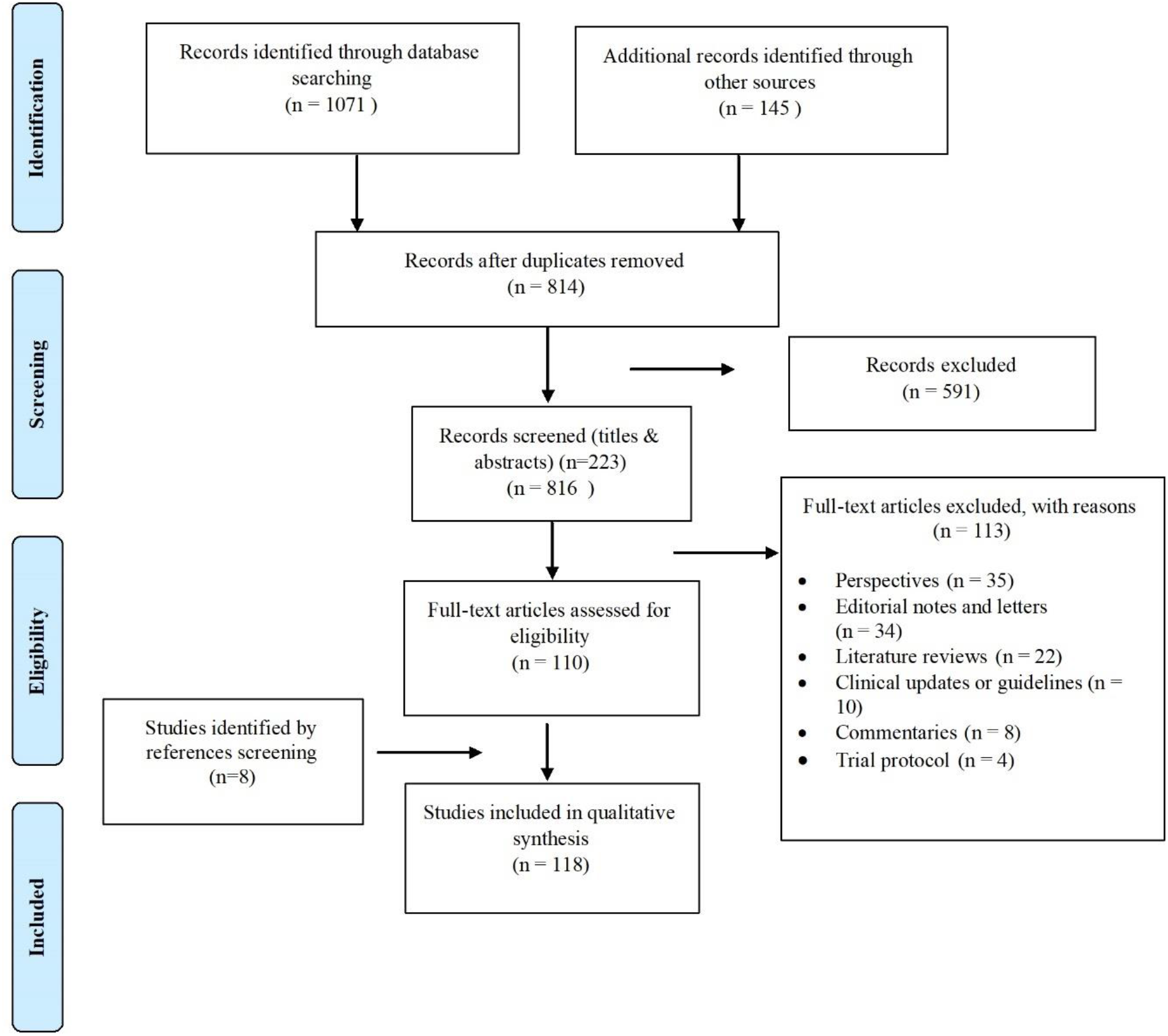
Prisma Chart.

### Description of included studies

Of 118 included studies, 59 (50%) were case series or case reports, 47 (39·8%) were observational studies based on hospital records and 7 (59%) of studies were randomised control trials. Most of the studies (61%) were conducted in low- and middle-income countries (LMICs), consonant with the trajectory of the pandemic at that stage, with the majority conducted in China - 61 (51·7%), followed by USA - 17 (14·4%), Italy - 11 (9·3%) and France - 7 (5·9%) respectively (Supplementary table 1). All the studies reported data of hospitalised COVID-19 patients. There were no reports of non-hospitalised COVID-19 patients in our results.

### Antibiotic prescribing and illness severity

Severity of illness was not reported in all studies. Just over half reported the severity of illness using four categories (severe, critical, moderate and mild) and the rest remainder used three groups (severe, moderate and mild). In order to explore the potential role of severity of illness in decisions regarding antibiotic prescribing, we grouped severity of illness into two broader categories: severe or critical, and moderate or mild. 2630 patients (41·9%) fell into the severe or critical group and 3649 patients (58·1%) into mild or moderate group.

In the included studies, 8501 out of 10 329 COVID-19 patients (82·3%) were prescribed antibiotics. There was little difference in the mean rates of antibiotic prescribing with 75·4% in severe or critical vs 75·1% in mild or moderate groups (Table 1).

**Table 1.**
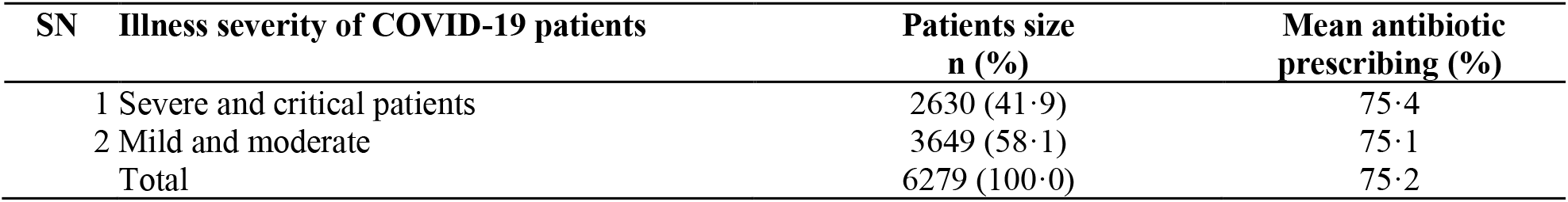
Severity of illness and antibiotic prescribing.

| SN | Illness severity of COVID-19 patients | Patients size<br>n (%) | Mean antibiotic<br>prescribing (%) |
| --- | --- | --- | --- |
| 1 | Severe and critical patients | 2630 (41.9) | 75.4 |
| 2 | Mild and moderate | 3649 (58.1) | 75.1 |
|  | Total | 6279 (100.0) | 75.2 |

### Antibiotic prescribing and health outcomes

We further explored the relationship between antibiotic prescribing and health outcomes (length of hospital stay (LOS), discharge rate and mortality rate (all these health outcomes were calculated at the time of publication of those studies; some patients were still in hospital and these patients were not included in their calculation). The results show that patient mortality was higher in studies for patients all given antibiotics compared to studies that majority of patients were not given antibiotics (26·5% vs 2·3%), LOS was longer (12·5 days vs 10·3 days), however, discharge rate was also higher (76·2% vs 73·2%) (Table 2).

**Table 2.**
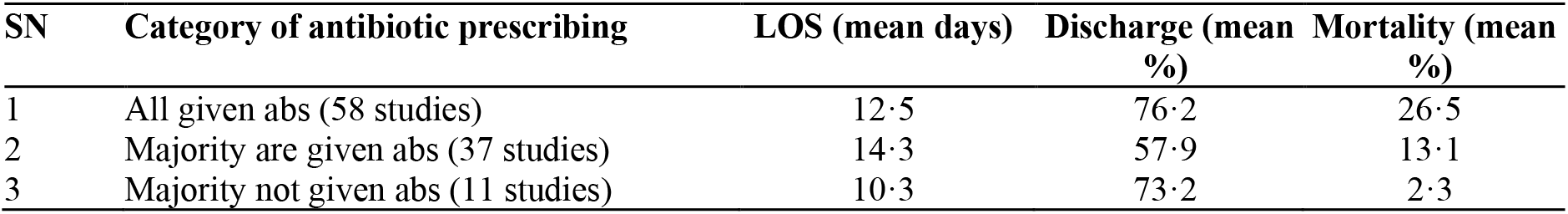
Antibiotic prescribing categories and outcomes.

### Frequently prescribed antibiotics

We extracted the details of prescribed antibiotics from all the included studies where this information was available. 33·9% of included studies did not report the details of antibiotics but only mentioned that antibiotics or empirical antibiotics were used in treatment.

Among the 78 studies that reported type of antibiotics used in the treatment of COVID-19 patients, (Fig 2) azithromycin (macrolides and ketolides) was the most frequently prescribed antibiotic (accounting for 28.0% of studies); followed by ceftriaxone (17·8%, 3^rd^ generation Cephalosporins), moxifloxacin (14·4%, quinolones and fluroquinolones), meropenem (14·4%, carbapenems and other penems), and Piperacillin/tazobactam (12·7%, antipseudomonal penicillins). It is not possible to tabulate prescribing percentages of these frequently prescribed antibiotics for the treatment of hospitalised COVID-19 patients as most studies, except case report or case series, did not report the percentage of each prescribed antibiotic in the treatment of COVID-19 patients. Notably, the frequently prescribed antibiotics are all broad-spectrum antibiotics.

**Fig 2.**
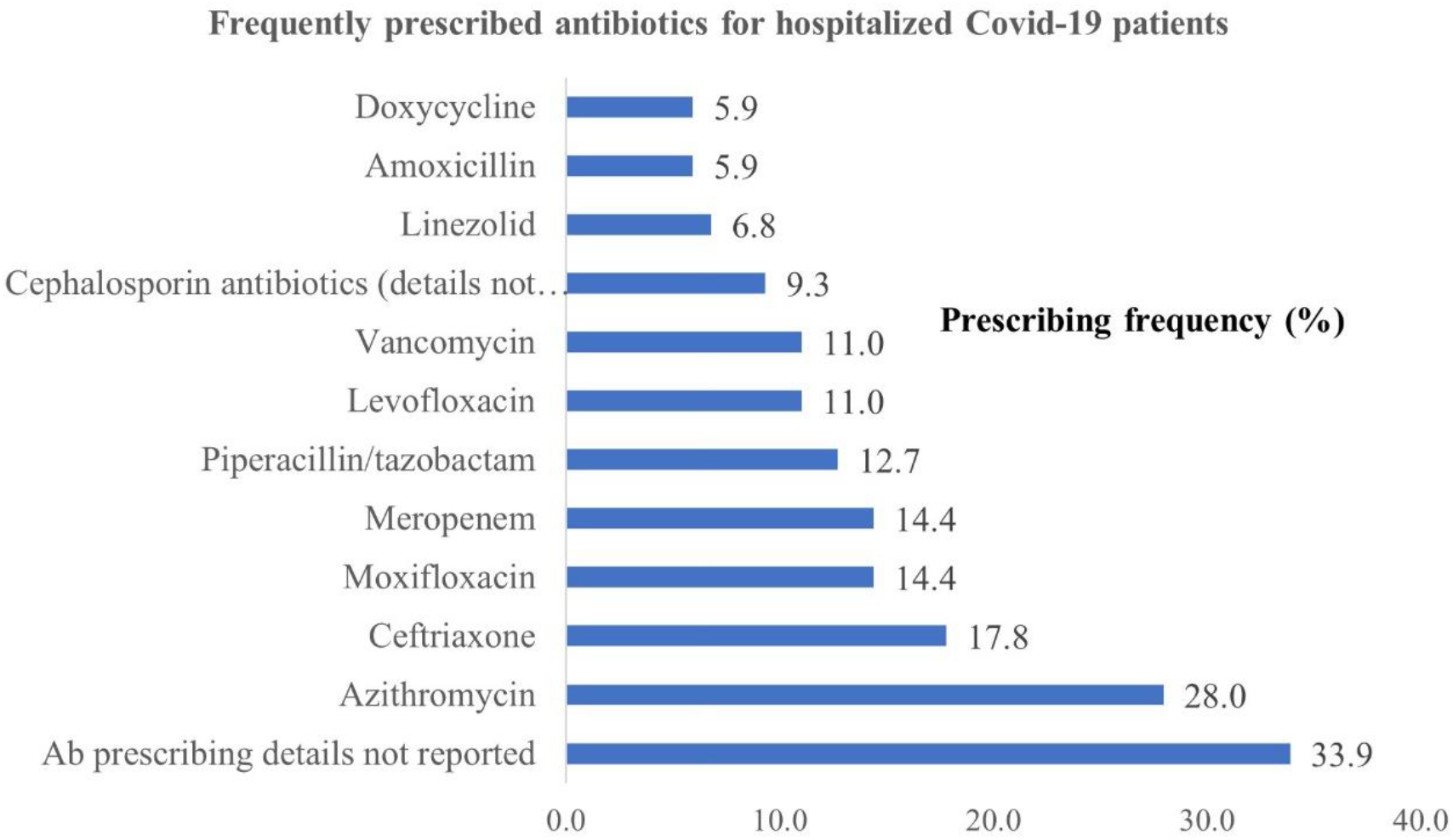
Frequently prescribed antibiotics for Covid-19 patients.

### Antibiotic prescribing scenarios

We summarized 20 different scenarios when antibiotics were prescribed based on the evidence available in our included studies (Table 3). We asked two experienced clinicians in infectious diseases with expertise in AMS practices to classify each antibiotic prescribing scenario as: 1) with clinical justifications; 2) without clinical justifications; 3) not sure. In addition to microbiological analysis; sepsis, elevated white blood cells or procalcitonin are also signs of bacterial infection and antibiotics prescribed under those circumstances were considered as “with clinical justifications”. There were some ambiguous cases (around 30% of scenarios) on which both experts found difficult to make judgements regarding whether antibiotics should be prescribed or not; we categorised those as “not sure”. A relatively high proportion (around 45%) of scenarios described in the included studies were categorised by both experts as “without clinical justifications”.

**Table 3.**
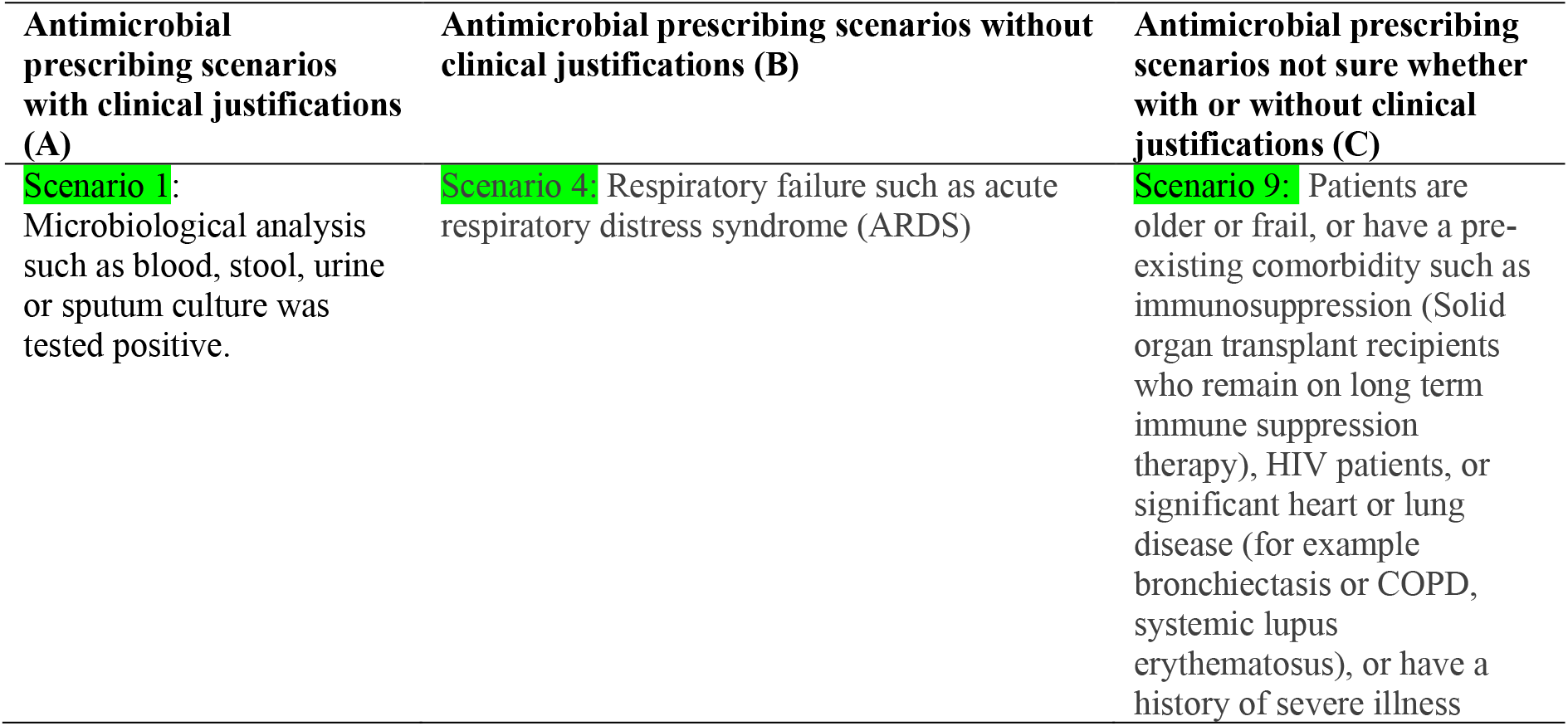
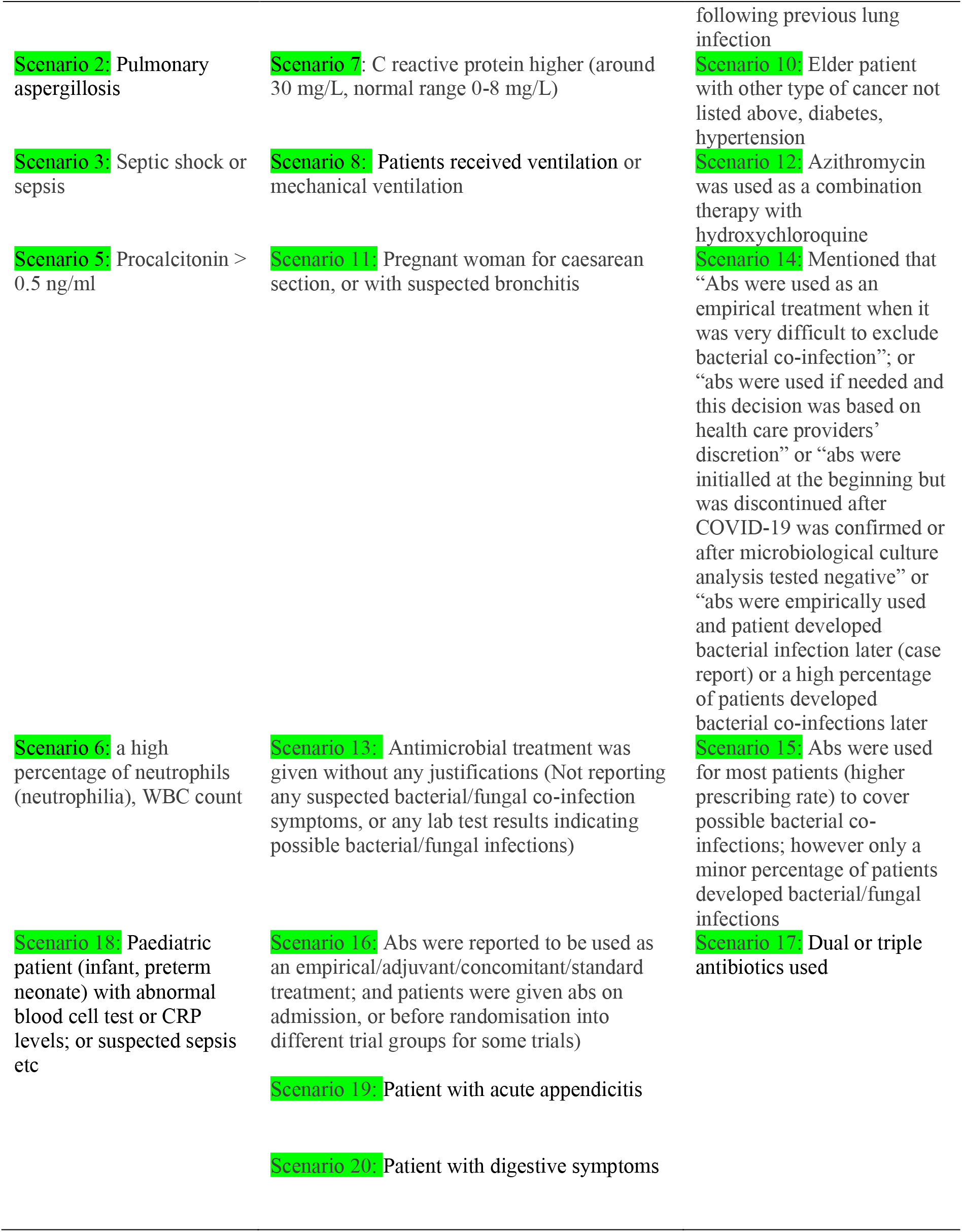
Antimicrobial prescribing scenarios of hospitalised COVID-19 patients.

We also categorized the frequency of each antibiotic prescribing scenario by illness severity (Supplementary table 2). We found that only 12·9% of severe or critically ill patients were prescribed antibiotics with stated clinical justifications, and this proportion was 13·6% for mild or moderate patients. There was no difference in the proportions of COVID-19 patients who were prescribed antibiotics without clinical justifications for mild or moderate patients as compared to severe or critical patients (51·5% vs 41·9%). Antibiotic prescribing scenarios classified as “not sure” were 45·2% for severe or critical patients as compared to 34·8% for mild or moderate patients.

### Severity of illness, antibiotic prescribing justifications and health outcomes

All studies with all severe or critical patients had a higher mortality rate than all studies with mild or moderate patients (53·1% vs 0·2%), similarly with lower discharge rate (96·2% vs 36·6%) and higher LOS (17·4% vs 8·7%) (Table 4). Table 5 shows relationship between antibiotic prescribing justifications and health outcomes. Mortality rate was lower for those patients who were provided antibiotics with clinical evidence of infections compared to those who were given without clinical justifications (9·5% vs 13·1%), discharge rate was higher (80·9% vs 69·3%) and LOS was lower (9·3 days vs 12·2 days). These results clearly show a relationship between evidence-based antibiotic prescribing and reduced mortality and length of hospital stay.

**Table 4.** Severity of illness and health outcomes.

| SN | Severity of illness (categories) | LOS (mean days) | Discharge (mean %) | Mortality (mean %) |
| --- | --- | --- | --- | --- |
| 1 | All severe/critical (16 studies) | 17.4 | 36.6 | 53.1 |
| 2 | Majority were severe/critical (4 studies) | 18.0 | 77.9 | 5.8 |
| 3 | Majority were mild/moderate (33 studies) | 12.0 | 60.5 | 4.8 |
| 4 | All mild/moderate (20 studies) | 8.7 | 96.2 | 0.2 |

**Table 5.**
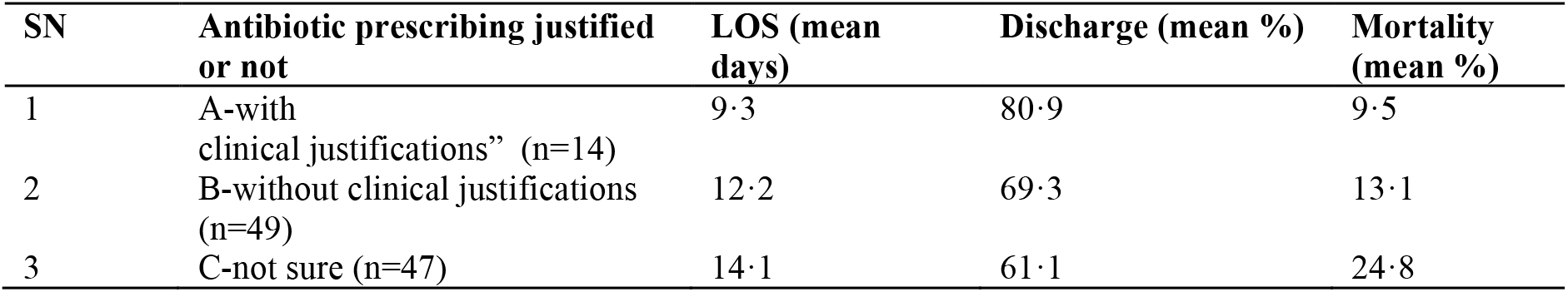
Antibiotic prescribing justifications and health outcomes.

### Secondary infections and health outcomes

Nine of the 118 studies specifically reported on secondary infections (5 from LMICs and 4 from HICs). Out of a total sample size of 820 in these studies, 74·4% patients had diagnosed secondary infections (n=610) and 51·3% of these patients were serious or critically ill (n=313). In perspective to our total sample size across all studies (6279 patients), the percentage of patients with confirmed or diagnosed secondary infections was 9·7%. Compared to total patients (n=6279), patients with secondary infections had higher mean LOS (20·4 days vs 12·4 days), lower discharge rate (54·8% vs 65·6%) and higher mortality rate (43·7% vs 16·3%) (Table 6).

**Table 6.** Secondary infections and health outcomes.

| Descriptions | Severe/critical n (%) | Mild/moderate n (%) | Mean length of stay (days) | Mean discharge rate (%) | Mean mortality rate (%) |
| --- | --- | --- | --- | --- | --- |
| Total patients with secondary infections (n=610) | 313 (51.3%) | 297 (48.7%) | 20.4 | 54.8 | 43.7 |
| Total sample size (N=6279) | 2630 (41.9%) | 3649 (58.1%) | 12.4 | 65.6 | 16.3 |

In total, 41 antibiotic prescriptions were issued across these nine studies. The five most frequently used antibiotics were piperacillin/tazobactam (14·6%), meropenem (9·8%), moxifloxacin (9·8%), ceftriaxone (7·3%), azithromycin (4·9%), linezolid (4·9%) and voriconazole (4·9%).

These studies reported 34 underlying conditions of the patients, with hypertension (14·7%, 5/34), diabetes (11·8%, 4/34), heart disease (11·8%, 4/34) and respiratory diseases (8·8%, 3/34) the top four underlying conditions (Supplementary Fig 1).

### Gender and Health Outcomes

Comparing the health outcomes of LOS, discharge rate and mortality rate by gender, we found that male patients compared to female patients had a higher mortality rate (37·7% vs 20%), lower discharge rate (75·2% vs 91·7%) and greater LOS (13·4 days vs 11·4 days) (Supplementary Table 3). These findings are in line with other recent studies conducted with COVID-19 patients across the world ^24,25^.

## Discussion

As the numbers of people with COVID-19 continue to increase globally, the widespread use of antibiotics for the treatment of COVID-19 patients and the potential consequences of this for antimicrobial resistance is a growing concern.

Our included studies discussed data of hospitalised patients; none of the studies discussed the data of non-hospitalised patients (e.g. care home patients, patients confined at home with mild symptoms). This is probably because this topic was not explored yet in the first six months of the pandemic. Data synthesis from 118 studies included in this review shows that around 82·3% of COVID-19 patients (8501 out of 10 329 patients) were prescribed antibiotics, whereas antibiotic prescribing percentages of COVID-19 patients were almost 100% in the early clinical reports from hospitals in Wuhan.^26-28^ It is unsurprising that doctors were giving antibiotics almost universally to treat this previously unknown respiratory infection at the beginning of the pandemic, while resort to antibiotics would be expected to decline with increasing knowledge about the novel coronavirus. More surprisingly, the antibiotic prescribing rate does not vary with illness severity. With 75·4% severe or critical patients being given antibiotics vs 75·1% of mild or moderate patients, although more seriously ill patients have a higher risk of developing secondary bacterial infections.

The high rate of unnecessary antibiotic prescribing for mild and moderate COVID-19 patients is alarming and inconsistent with WHO COVID ^29^ and UK NICE guidelines ^30^. The unnecessary over-prescription of antibiotics for mild and moderate COVID-19 patients, not only indicates the emerging challenges of antimicrobial stewardship practice in hospitals around the world at least in the early phase of the pandemic but suggests a potential increase in resistant bacterial pathogens and antimicrobial resistance in affected countries in the coming months.

Moreover, the five most frequently prescribed antimicrobials (azithromycin, ceftriaxone, moxifloxacin, meropenem, piperacillin/tazobactam) are all classified as critically important antimicrobials (CIA) for human medicine.^31^ Persistent use of those critically important antibiotics will provoke the emergence of MDR strains and a decline in the effectiveness of these compounds,^32^ which poses a threat to survival rates from serious infections, neonatal sepsis and hospital infections, thus limiting the potential health benefits of surgery, transplants and cancer treatment.^33^ A systematic review has also provided evidence on the emergence of antimicrobial resistance after mass azithromycin distribution for trachoma control programmes in Sub-Saharan Africa; macrolide resistance after azithromycin distribution was reported in three of the five organisms studied and there was little evidence for absence of resistance in *Chlamydia trachomatis* after azithromycin treatment, suggesting that Azithromycin may not remain effective for trachoma programmes.^34^ The impact of widespread use of ceftriaxone on development of resistance to third generation cephalosporins among clinical strains of Enterobacteriaceae and other non-enteric bacteria is already well known.^35-37^ If resistance to azithromycin, ceftriaxone and other broad-spectrum antibiotics becomes widespread due to their massive use during the pandemic, there would be very few alternative antibiotics available in the market and these alternatives antibiotics are likely to be unaffordable for the majority of patients especially in low and middle-incomes countries.

Clinicians were under extreme pressure in the treatment of COVID-19 patients especially in the early phase of the pandemic. It is increasingly well recognised that COVID-19-related concerns or unknowns may change prescription behaviours of healthcare professionals (HCPs) and drive antibiotic use.^38^ HCPs were eager to save the life of patients by trying different existing drugs including antibiotics. For example, azithromycin was frequently used in combination with hydroxychloroquine as a treatment regime in many clinical studies, although this has since been confirmed in large international trials as not effective.^39^ Also, the gold standard diagnostic confirmation of secondary bacterial/fungal infections through microbiological culturing is costly, time consuming and not available in smaller hospitals, making rapid exclusion of secondary infections very difficult and pushing HCPs to error on the side of treating with antibiotics both as directed and empirical therapy.

We would expect adherence to established antibiotic stewardship programmes (ASP) to decline as HCPs struggle to save the lives of patients with COVID-19. Nonetheless, the low proportion of antibiotic prescribing (less than 14%) for COVID-19 patients with a clinical justification for assuming the presence of bacterial infection is surprising. Around 45% of antibiotic prescribing scenarios described in the reviewed studies were classified as “not sure” for severe or critical patients, but only around 35% for mild or moderate patients, suggesting that in cases of greater severity, HCPs are more likely to resort to antibiotic treatment even in the absence of clinical indications suggestive of bacterial infection. Overall, 40-50% of antibiotic prescribing (41·9 % for severe or critical patients vs 51·5% for mild or moderate patients) in the included studies occurred without clinical indications of bacterial infection. Only 9·7 % of the total number of COVID-19 patients in the reviewed studies was reported to have a secondary bacterial infection, consistent with findings from other publications.^40,41^Among the three most common prescribing scenarios reported in our included studies (Supplementary table 3: scenarios 13, 14 & 16), two depicted unwarranted antibiotic prescribing: 1) without reporting any suspected bacterial/fungal infections or any lab test results indicating possible bacterial/fungal infections (scenario 13); 2) antibiotics were reported to be used as an empirical/adjuvant/concomitant/standard/prophylactic treatment; and patients were given antibiotics on admission immediately, or before randomisation into different treatment groups in some trials (scenario 16)

Our analysis also provides some evidence regarding the effect of antibiotic use on treatment outcomes, as measured by LOS, discharge rate and mortality rate of hospitalised COVID-19 patients. We found that overall, patients given antibiotics had a higher mortality rate (26·5% vs 2·3%) and longer LOS (12·5 days vs 10·3 days) than, but similar discharge rates (76·2% vs 73·2%), to, the majority of patients who were not given antibiotics. We would expect greater use of antibiotics to be associated with worse outcomes due to severe and critical COVID-19 patients being more likely to receive antibiotics, but this is not supported by our finding that antibiotic prescribing was not associated with illness severity. Our results suggest that antibiotic treatment may not improve COVID-19 patients’ treatment outcomes. Patients who were prescribed antibiotics with suspected bacterial infection were also more likely to have negative clinical outcomes, including higher mean LOS (20·4 days vs 12·4 days), lower discharge rate (54·8% vs 65·6%) and higher mortality rate (43·7% vs 16·3%) compared to mean patient treatment outcomes. This suggests that COVID-19 patients with secondary infections may not gain additional benefit from antibiotic treatment for these secondary infections. This is consonant with findings from a recent clinical study exploring the treatment outcomes of 1123 COVID-19 patients from Wuhan that compared antibiotics treatment between patients with suspected bacterial infection as compared to those with no evidence of bacterial infection, which found that antibiotic therapy was associated with increased mortality and most patients would not benefit from antibiotics treatment. Intravenous moxifloxacin and meropenem actually increased the mortality of patients with suspected bacterial infection, and penicillin and meropenem treatment was associated with increased mortality in patients with no evidence of bacterial infection.^42^ Another retrospective study that reviewed the medical charts of 48 intubated ICU patients admitted between Apr and May 2020 in Switzerland, reported that early administered antibiotics do not appear to significantly impact mortality or delay hospital-acquired infections in critically ill COVID-19 patients. ^43^ Large multi-centre studies are urgently needed to provide direct evidence for our findings and further investigate the impact of antibiotic treatment on mortality and other treatment outcomes of COVID-19 patients with different severities. Although empirical, adjuvant and prophylactic use of antibiotics for severe and critical patients that is still endorsed in WHO, UK & China guidelines,^29,30,44^ may not bring expect benefits.

Strengths of the review include coverage of 118 studies across a wide range of study designs published during the six-month review period, giving a full landscape of antibiotic use trends for Covid-19 patients during the first wave of the pandemic. Likewise, we included all published literature in English and Chinese. Although more is now known about COVID-19 and effective treatments, as it spreads across the globe there is no reason to believe that antibiotics are not still being used widely, particularly in more recently affected low and middle-income countries with limited resources, so our findings continue to have relevance. This review and evidence synthesis also has several limitations. First, our search included five databases only. However, the inclusion of English and Chinese literature and search of reference list of eligible studies ensured that we were able to identify the majority of published studies. Due to the heterogeneity of the studies, meta-analysis was not conducted. However, the inclusion of a wide range of study designs may help in providing a complete picture of the global antibiotic prescribing patterns during the COVID-19 pandemic, which is lacking in the current scientific literature.

## Conclusions

This review and evidence synthesis demonstrates that during the first six months of the COVID-19 pandemic, antibiotic prescribing in hospitals was not associated with illness severity. Most Covid-19 patients with mild or moderate illness, who should not receive antibiotics according to international guidelines, had been prescribed antibiotics in the reports and studies we reviewed. We also find that antibiotics were frequently prescribed without clinical or microbiological indications of bacterial infection.

The evidence reviewed suggests that even where secondary bacterial infection is present, antibiotic prescribing may not improve treatment outcomes in COVID-19 patients. Until more clinical data become available to verify these findings, considerable caution is warranted when considering antibiotic treatment in COVID-19 cases, even for severe and critically ill patients. Thus the widespread use of antibiotics for COVID-19 may not only magnify the problem of antibiotic resistance globally and render currently available antibiotics ineffective, but also provide little or no benefit for COVID-19 patients. A further rapidly review to capture changes of antibiotic prescribing and secondary bacterial infections in subsequent phases of the pandemic is currently underway.

## Data Availability

Raw data could be available upon request

## Acknowledgments

The authors acknowledge MRC & Newton Fund support through a UK-China AMR Partnership Hub award (MR/S013717/1). We would like to acknowledge Prof Paul Little (School of Medicine, University of Southampton, Southampton, UK) and Prof Bo Zheng (Institute of Clinical Pharmacology, Peking University First Hospital, Beijing, China) for their contributions to reviewing and refining the clinical justifications on antibiotic prescribing scenarios of hospitalised COVID-19 patients in this review.

## Author Contributions

HL, WC, NP, NA and HW were responsible for developing the research questions and the study design. WC, NA and HW were responsible for literature search and data extraction; NP & WC were responsible for data analysis. WC, NA, NP, HW, GY and HL drafted the paper. All authors read, commented on and approved the final manuscript.

---

**Supplementary Fig 1.**
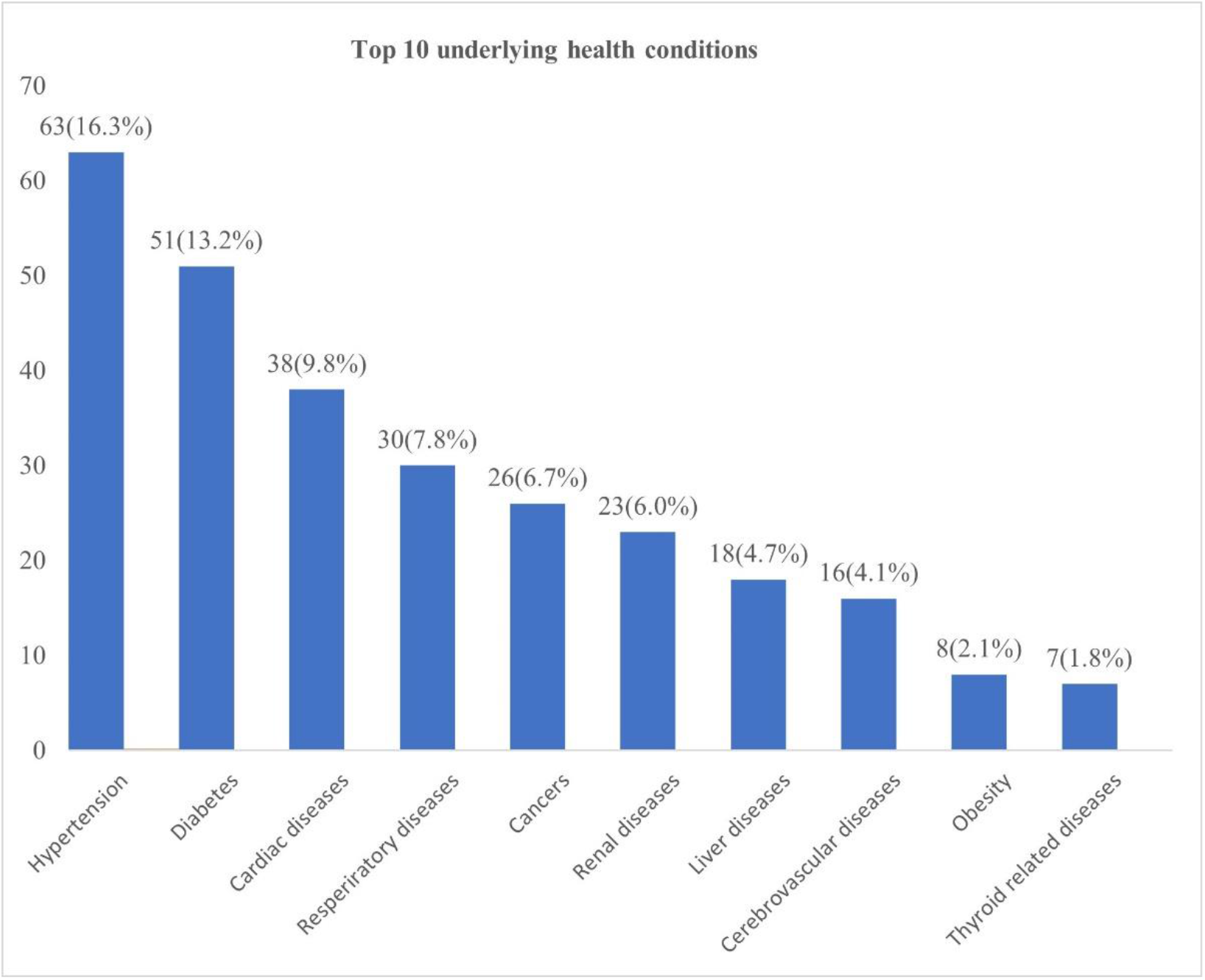
Top 10 underlying health conditions of Covid-19 patients.

**Supplementary Table Legends**

Supplementary Table 1 Description of included studies

Supplementary Table 2. Categories of illness and antibiotics prescribing

Supplementary Table 3. Severity of illness and scenario of antibiotic prescribing

Supplementary Table 4. Types of study based on gender and health outcomes (LOS, discharge and mortality)

**Supplementary Table 1.** Description of included studies

| SN | Description | Frequency (n) | Percentage (%) |
| --- | --- | --- | --- |
| **Types of studies** |  |  |  |
| 1 | Case reports/case series | 59 | 50.0 |
| 2 | RCT | 7 | 5.9 |
| 3 | Cohort | 5 | 4.2 |
| 4 | Other (descriptive and other studies) | 47 | 39.8 |
|  | Total | 118 | 100 |
| **Study countries or areas** |  |  |  |
| 1 | LMICs (China, Iran, Bhutan, Brazil, Colombia, Iraq, Nepal, Philippines, Uganda, Vietnam) | 72 | 61.0 |
| 2 | HICs (USA, Italy, France, Spain, United Kingdom, Belgium, Hong Kong (China), Japan, Poland, South Korea) | 56 | 39.0 |
|  | Total | 118 | 100 |

**Supplementary Table 2.** Severity of illness and scenario of antibiotic prescribing

| SN | Scenario of antibiotics prescribing | Severe and critical illness and scenario of antibiotic prescribing |  | Mild and moderate illness and scenario of antibiotic prescribing |  |
| --- | --- | --- | --- | --- | --- |
|  |  | Frequencies (n) | Percentage (%) | Frequencies (n) | Percentage (%) |
| 1 | Scenario 1 (A) | 3 | 4.8 | 2 | 3.0 |
| 2 | Scenario 4 (B) | 2 | 3.2 | 2 | 3.0 |
| 3 | **Scenario 6 (A)** | 4 | 6.5 | 6 | 9.1 |
| 4 | Scenario 7 (B) | 2 | 3.2 | 4 | 6.1 |
| 5 | Scenario 8 (B) | 2 | 3.2 | 5 | 7.6 |
| 6 | **Scenario 9 (C)** | 8 | 12.9 | 2 | 3.0 |
| 7 | Scenario 10 (C) | 2 | 3.2 | 2 | 3.0 |
| 8 | **Scenario 12 (C)** | 4 | 6.5 | 5 | 7.6 |
| 9 | **Scenario 13 (B)** | 8 | 12.9 | 13 | 19.7 |
| 10 | **Scenario 14 (C)** | 11 | 17.7 | 10 | 15.2 |
| 11 | Scenario 15 (C) | 2 | 3.2 | 2 | 3.0 |
| 12 | **Scenario 16 (B)** | 11 | 17.7 | 9 | 13.6 |
| 13 | Scenario 17 (C) | 1 | 1.6 | 2 | 3.0 |
| 14 | Scenario 18 (A) | 1 | 1.6 | 1 | 1.5 |
| 15 | Scenario 20 (B) | 1 | 1.6 | 1 | 1.5 |
| Total |  | 62 | 100 | 66 | 100 |

[Note: every study may have more than one abs prescribing scenario; A- with clinical justifications; B – without clinical justifications; C – not sure]

**Supplementary Table 3.** Types of study based on gender and health outcomes (LOS, discharge and mortality)

| SN | Gender breakdown of studies | LOS (mean days) | Discharge (mean %) | Mortality (mean %) |
| --- | --- | --- | --- | --- |
| 1 | Studies with female patients only (18 studies) | 11.4 | 91.7 | 20.0 |
| 2 | Studies with male patients only (27 studies) | 13.4 | 75.2 | 37.5 |
| 3 | Studies with mix of male and female patients (72 studies) | 13.1 | 61.5 | 12.6 |

[Note: 1 study did not provide required information]

